# A Robust and Reproducible Connectome Fingerprint of Ketamine is Highly Associated with the Connectomic Signature of Antidepressants

**DOI:** 10.1101/2020.04.10.20061085

**Authors:** Chadi G. Abdallah, Kyung-Heup Ahn, Lynnette A. Averill, Samaneh Nemati, Christopher L. Averill, Samar Fouda, Mohini Ranganathan, Peter T. Morgan, Deepak C. D’Souza, Daniel H. Mathalon, John H. Krystal, Naomi R. Driesen

## Abstract

Over the past decade, various N-Methyl-D-Aspartate modulators have failed in clinical trials, underscoring the challenges of developing novel rapid-acting antidepressants based solely on the receptor or regional targets of ketamine. Thus, identifying the effect of ketamine on the brain circuitry and networks is becoming increasingly critical. In this longitudinal functional magnetic resonance imaging study of data from 265 participants, we used a validated predictive model approach that allows the full assessment of brain functional connectivity, without the need for seed selection or connectivity summaries. First, we identified a connectome fingerprint (CFP) in healthy participants (Cohort A, n=25) during intravenous infusion of a subanesthetic dose of ketamine, compared to normal saline. We then demonstrated the robustness and reproducibility of the discovered Ketamine CFP in two separate healthy samples (Cohort B, n=22; Cohort C, n=18). Finally, we investigated the Ketamine CFP connectivity at 1-week post treatment in major depressive disorder patients randomized to 8 weeks of sertraline or placebo (Cohort D, n=200). We found a significant, robust, and reproducible Ketamine CFP, consistent with reduced connectivity within the primary cortices and within the executive network, but increased connectivity between the executive network and the rest of the brain. Compared to placebo, the Ketamine CFP connectivity changes at 1-week predicted response to sertraline at 8-weeks. In each of Cohort A-C, ketamine significantly increased connectivity in a previously identified Antidepressant CFP. Investigating the brain connectivity networks, we successfully identified a robust and reproducible ketamine biomarker that is related to the mechanisms of antidepressants.

**Graphical Abstract:** 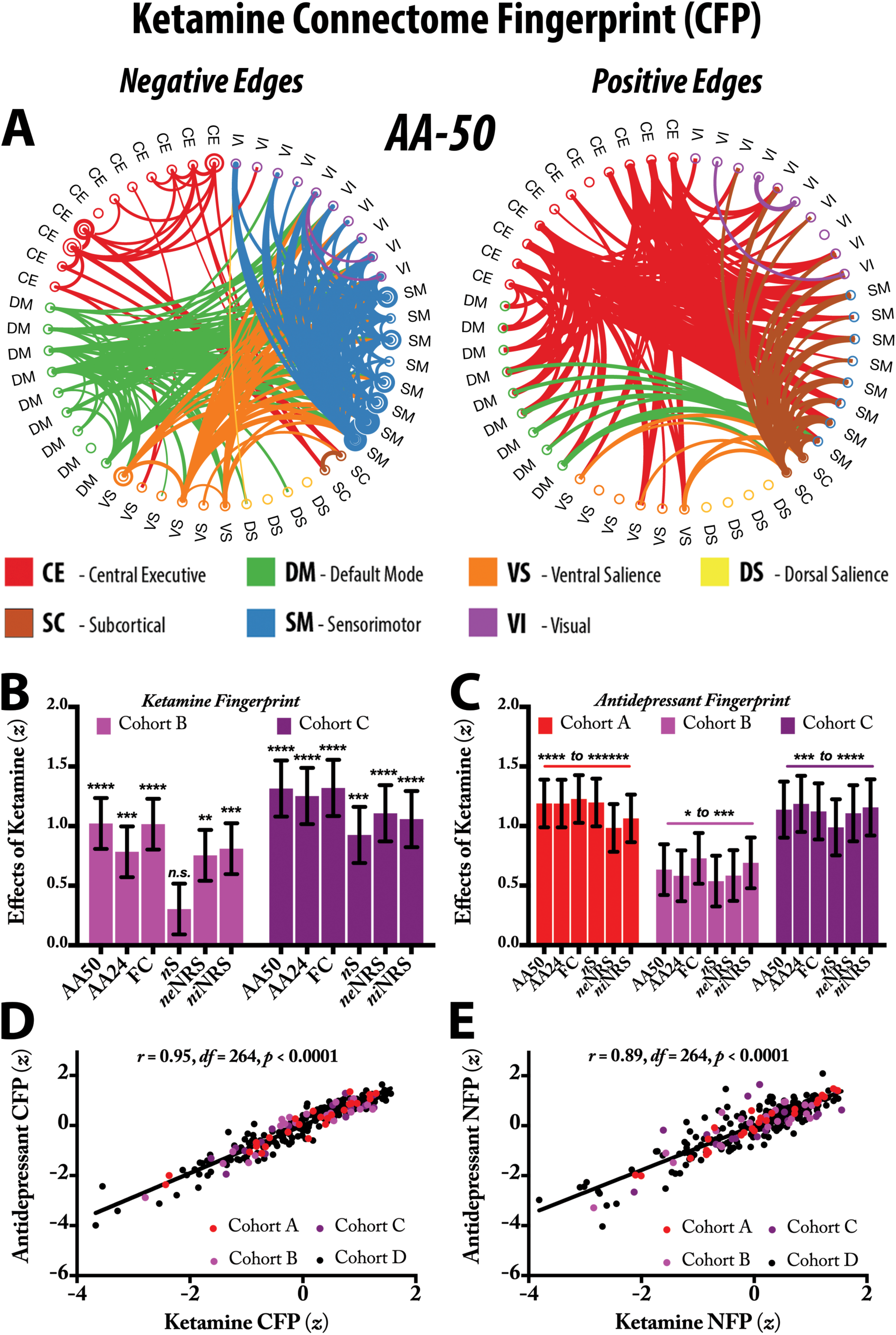

## INTRODUCTION

Major depressive disorder (MDD) remains among the most disabling illnesses, with increased risk of suicide and high treatment resistance. Moreover, most standard MDD treatments are slow acting, requiring weeks to months to achieve full therapeutic effects. The discovery of the rapid-acting antidepressant (RAAD) effects of ketamine has generated excitement and optimism in the field [1-3]. Over the past two decades, there has been a longstanding desire to identify novel antidepressants that preserve the RAAD effects of ketamine, without its side effects and abuse liability [4]. Molecular and cellular hypotheses related to the mechanism of ketamine efficacy suggest a number of novel directions to pursue in RAAD drug development [5]. However, broadening the perspective to include ketamine’s impact on brain functional connectivity networks may enrich the effort to identify novel treatment mechanisms. In particular, a network view of ketamine’s effects may help us to better understand the underlying mechanisms, supplementing current knowledge of ketamine’s neurochemical properties and regional brain targets.

Profiling the connectivity of all the brain’s networks, a process termed functional connectome fingerprinting, has been successfully applied to predict behavior (e.g., [6]) and recently identified a unique connectome fingerprint (CFP) that predates and predicts response to antidepressants [7]. In the current study, we employed this predictive model approach to identify robust and reproducible functional connectivity signatures of ketamine. This approach is a combination of the network restricted strength (NRS) methods and the connectome-based predictive modeling [8, 9]. The connectome-based predictive model is a series of analytic steps that selects and summarizes features, builds and applies models, and finally assesses their predictive significance [9]. Importantly, this predictive model retains the ability to back-translate findings to the original feature space, a unique advantage that is often lacking in many machine learning approaches [6, 9]. In the NRS approach, we use whole-brain parcellation along with the Akiki-Abdallah (AA) hierarchical connectivity atlas [7, 10] to significantly reduce the number of edges, while retaining the upstream canonical intrinsic network affiliations to the central executive (CE), default mode (DM), ventral salience (VS), dorsal salience (DS), subcortical (SC), sensorimotor (SM) and visual (VI) networks [11]. The main goal of this network restricted strength predictive model (NRS- PM) is not to assess the strength of predictions, but rather to identify functional connectivity signature that is associated, significantly and consistently, with the outcome of interest (e.g., ketamine effects) [11]. This approach has many strengths, including: (1) the predictive model design may enhance reproducibility by providing protection against overfitting, which is an issue with traditional interpretive analyses; (2) the multivariate pattern analysis allows the full assessment of the connectome, without the inherent increase of Type I error due to univariate multiple comparisons or the need to limit the analysis to a few seeds; and (3) the results are, by design, network-based that both informs the neurobiological models and facilitates the integration of findings.

In biomarker development, it is essential that the putative marker is: (1) *robust*, having a large effect size so it is evident even in relatively small samples; (2) *reproducible*, being stable within a population and generalizing to new observations; and (3) *relevant*, associated with the clinical outcome of interest such as predicting group differences or changes in clinical status or being responsible for these changes [12]. In this study, our primary aim was to evaluate the robustness and reproducibility of the effects of ketamine on human cortical functional connectivity (here termed, “Ketamine CFP”) across three samples of healthy participants (Cohort A, B and C; n=18-25). As a secondary aim, we explored the clinical relevance of the Ketamine CFP in a randomized trial of patients with MDD (Cohort D; n=200). We hypothesized that the NRS-PM in Cohort A would identify a significant CFP of the effects of ketamine, and that this CFP would be robust and reproducible in Cohort B and C. We also investigated, in Cohort D, whether the Ketamine CFP connectivity predicts the antidepressant response in MDD patients treated with sertraline, compared to placebo. Finally, to assess the brain localization of ketamine’s signature, we analyzed changes in connectivity strength of all nodes, termed the nodal fingerprint (NFP) [11].

## METHODS

### Participants and Procedures

The data (n=265) used in this study were collected as part of 4 independent trials [13-16]. Functional connectivity analyses were not performed previously in data collected in the primary sample (Cohort A). Therefore, the ketamine connectivity signatures were identified in Cohort A [14], while Cohort B, C, and D were used to assess robustness, reproducibility, and clinical relevance [13, 15, 16]. All studies were approved by Institutional Review Boards, and all participants signed informed consents.

Full details of the trials A-D were reported previously [13-16]. Of relevance to the current report, Cohort A (n=25; mean age *±SEM* = 30 ±1.9 years; 19 men), Cohort B (n=22; mean age *±SEM* = 29 ±1.5 years; 14 men), and Cohort C (n=18; mean age *±SEM* = 28 ±0.9 years; 18 men) included healthy subjects who received brain functional magnetic resonance imaging (*f*MRI), during administration of intravenous normal saline then ketamine as an intravenous bolus (0.23 mg/kg) followed by an intravenous infusion of 0.58 mg/kg throughout the scan. Participants were excluded if they had an unstable medical illness, a psychiatric disorder, or an MR contraindication. Cohort D (n=200; mean age *±SEM* = 38 ±0.9 years; 67 men) included unmedicated non-psychotic patients with MDD who were randomized to an 8-week course of up to 200 mg daily sertraline or to placebo (Table S1). Participants received *f*MRI scans at baseline and at Week 1 of treatment [11, 16]. Depression severity was rated on the 17-item Hamilton Depression Rating Scale (HAMD) at baseline and at Week 8 after treatment.

### Neuroimaging Acquisition and Processing

In the ketamine studies, *f*MRI scans were acquired during intravenous infusions of normal saline and then during ketamine, while subjects performed 8 spatial working memory runs (voxel size = 3.1×3.1×5mm^3^; TR=1500 ms, 166 frames; Cohort A/B) or 3 visual oddball runs (voxel size = 3.4×3.4×4.5mm^3^; TR=2000 ms, 210 frames; Cohort C) [13-15]. In consideration of reports of time effects [17], functional connectivity measures were computed on the early runs (i.e., ∼12 minutes) during each intravenous infusion of ketamine and of normal saline. In the MDD study, *f*MRI scans were acquired at rest (3.2×3.2×3.1mm^3^; TR=2000 ms; ∼12 min; baseline and Week 1 sessions) [7, 11, 18]. All studies included high-resolution structural MRI. Brain scans from all studies underwent the same surface-based preprocessing, using a pipeline adapted from the human connectome project (HCP; https://github.com/Washington-University/HCPpipelines) [19], as reported elsewhere [7, 11, 20-22]. Briefly, the preprocessing pipeline included FreeSurfer parcellation of structural scans, slice timing correction, motion correction, intensity normalization, brain masking, and registration of *f*MRI images to structural MRI and standard template. Then, the cortical gray matter ribbon voxels and each subcortical parcel were projected to a standard Connectivity Informatics Technology Initiative (CIFTI) 2mm grayordinate space (i.e., gray matter area in CIFTI). ICA-FIX was run to identify and remove artifacts [23, 24], followed by mean grayordinate time series regression (MGTR).

### Connectome and Nodal Predive Models

Full details of the network restricted strength (NRS) methods were previously reported [8, 11]. Briefly, the A424 atlas is used to segment the whole-brain gray matter into 424 nodes, and to compute average time series within each node [11, 25-27]. The Akiki-Abdallah hierarchical connectivity at 50 modules (AA-50; Fig. S1), 24 modules (AA-24; Fig. S2), and 7 modules (AA-7) were used to determine the network affiliation of the A424 nodes (https://github.com/emergelab). The full connectome is the Fisher-z transformation of the pairwise correlation coefficients. NRS connectome is the pairwise average connectivity of all modules at AA-50 and AA-24 [11]. Nodal strength (nS) is the average connectivity of a node to all other nodes. Nodal internal NRS (niNRS) is the average connectivity between each node and all other nodes within the same canonical connectivity network (i.e., AA-7). Nodal external NRS (neNRS) is the average connectivity between each node and all other nodes outside its canonical connectivity network [11]. The predictive models used were adapted from the connectome-based predictive model approach [9], as previously detailed [11]. The modeling includes feature selection in training subsamples, followed by fitting a linear predictive model, then applying the model to the test subsample [9]. As previously established, we used 1000 iterations of ten-fold cross-validation (CV) to ensure the stability of the models and to determine the statistical significance; that is by comparing true and random predictions [11].

### Statistical analyses

Descriptive statistics were calculated prior to statistical analysis. Data distributions were checked using normal probability plots. The statistical significance threshold was set at 0.05 (2-tailed tests). MATLAB (2018a; Mathworks Inc.) and the Statistical Package for the Social Sciences (version 24; IBM) software were used for the analyses. False Discovery Rate (FDR; *q* < 0.05) was used to correct for multiple comparisons. The improvement-by-treatment interaction was computed as the percent improvement in depression score multiplied by the treatment contrast (i.e., 1 for study drug and –1 for placebo control) [11]. NRS-PM at AA-50 is considered the primary CFP analysis, with two secondary CFPs at AA-24 and full connectome. The goal of the secondary CFPs is to inspect the upstream modules (i.e., AA-24) and to test the model without network restrictions (i.e., full connectome) [11]. Similarly, nS predictive model is considered the primary nodal fingerprint analysis, with niNRS and neNRS as secondary analyses to interrogate the shifts within and between networks, which are not captured by nS [11]. Connectivity per fingerprint was computed by multiplying the connectivity features (e.g., NRS at AA50) by the corresponding weighted fingerprint masks (e.g., the Antidepressant CFP from [11]). Paired *t-*tests were used to examine the effects of ketamine on the connectivity fingerprints. Pearson correlations were used to examine the association among connectivity fingerprints and between ketamine fingerprints and depression improvement following sertraline, compared to placebo. The study atlases, codes, and predictive models will be made publicly available at https://github.com/emergelab.

## RESULTS

### Ketamine Connectome Fingerprint (CFP)

The primary analysis, at AA-50, identified a CFP that is significantly associated with ketamine infusion, compared to normal saline (*r* = 0.66, *CV* = 10, *iterations* = 1000, *p* < 0.001). Secondary analyses also identified significant CFPs at AA-24 (*r* = 0.60, *CV* = 10, *iterations* = 1000, *p* < 0.001, *q* < 0.05) and at the full connectome (*r* = 0.65, *CV* = 10, *iterations* = 1000, *p* < 0.001, *q* < 0.05). As shown in Fig. 1, ketamine infusion was associated with reduced connectivity within the primary cortices (sensorimotor and visual) and within the central-executive-subcortical modules, but increased connectivity between the central-executive-subcortical and the rest of the brain. For the default mode and ventral salience modules, there is a shift from connectivity with primary cortices during saline (i.e., negative predictive edges) toward increased connectivity with the central-executive-subcortical modules during ketamine infusion (i.e., positive predictive edges) (Fig. 1).

**Figure 1.**
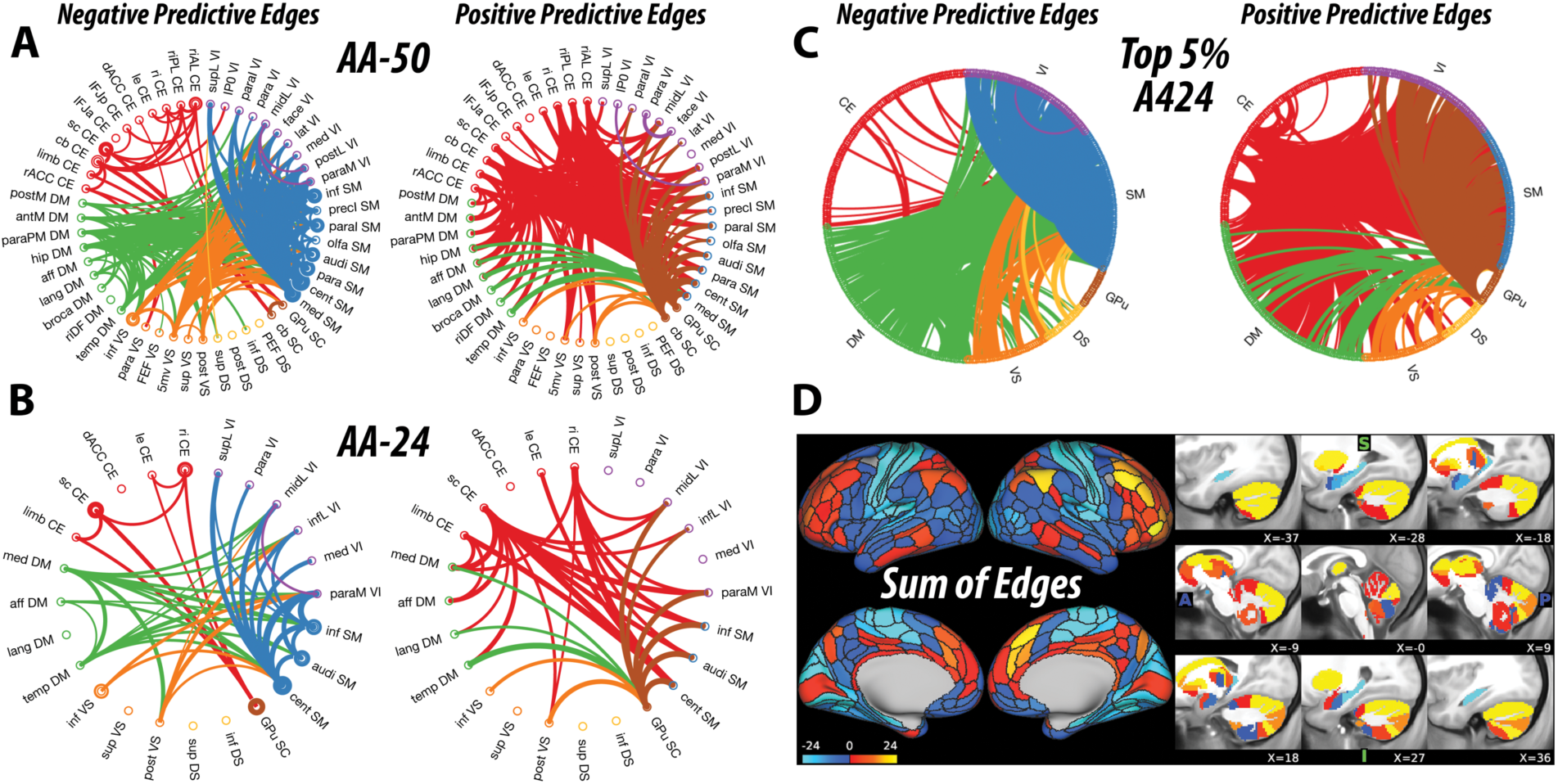
Ketamine Connectome Fingerprint (CFP). **A-C**. The circular graphs are labeled based on the Akiki-Abdallah (AA) whole-brain architecture at 50 modules (AA-50; primary CFP), 24 modules (AA-24), and the full connectome with 424 nodes (A424). Modules and nodes are colored according to their affiliation to the 7 canonical connectivity networks: central executive (CE), default mode (DM), ventral salience (VS), dorsal salience (DS), subcortical (SC), sensorimotor (SM), and visual (VI). Edges are colored based on the initiating module using a counter-clockwise path starting at 12 o’clock. Internal edges (i.e., within module) are depicted as outer circles around the corresponding module. Thickness of edges reflect their corresponding weight in the predictive model. The module abbreviations of AA-24 and AA-50, along with further details about the affiliation of each node are available at https://github.com/emergelab/hierarchical-brain-networks/blob/master/brainmaps/AA-AAc_main_maps.csv. Only edges of significant predictive models following correction are shown (all *p* ≤ 0.001). **C**. For the full connectome, it is not possible to visually discern the underlying signature considering the large number of edges retained. Therefore, as in previous studies, the circular graph is thresholded using nodal strength within the full connectome fingerprint as cutoff to retain the highest top 2.5% negative predictive edges and top 2.5% positive predictive edges. **D**. Shows the nodal degree of the full connectome fingerprint edges without a threshold. The color bar unit is arbitrary, reflecting the sum of weighted edges. All predictive models will be made publicly available at https://github.com/emergelab.

Due to the large number of predictive edges, visualizing the full connectome often yields undiscernible CFP [11]. However, inspecting the nodes with highest degree (i.e., top 2.5% of each of positive and negative predictive edges) revealed a pattern of connectivity signature comparable to the AA-50 CFP (Fig. 1C). The sum of edges of the full connectome model depicts the nodal degree of connectivity changes during ketamine, compared to normal saline. Note that only edges that were statistically significant in the model were depicted in the figures and included in the sum of edges.

### Ketamine Nodal Fingerprint (NFP)

The primary analysis, based on nS, identified an NFP that is significantly associated with ketamine infusion, compared to normal saline (*r* = 0.60, *CV* = 10, *iterations* = 1000, *p* < 0.001). Secondary analyses also identified significant NFPs based on niNRS (*r* = 0.61, *CV* = 10, *iterations* = 1000, *p* < 0.001, *q* < 0.05) and neNRS (*r* = 0.65, *CV* = 10, *iterations* = 1000, *p* < 0.001, *q* < 0.05). As shown in Fig. 2, ketamine infusion was associated with significant reduction in nS in the primary cortices and the hippocampus, but increased nS in the central-executive-subcortical regions. Inspection of the niNRS/neNRS results revealed a shift within the central-executive-subcortical networks, with reduced internal but increased external connectivity during ketamine infusion (Fig. 2C-D).

**Figure 2.**
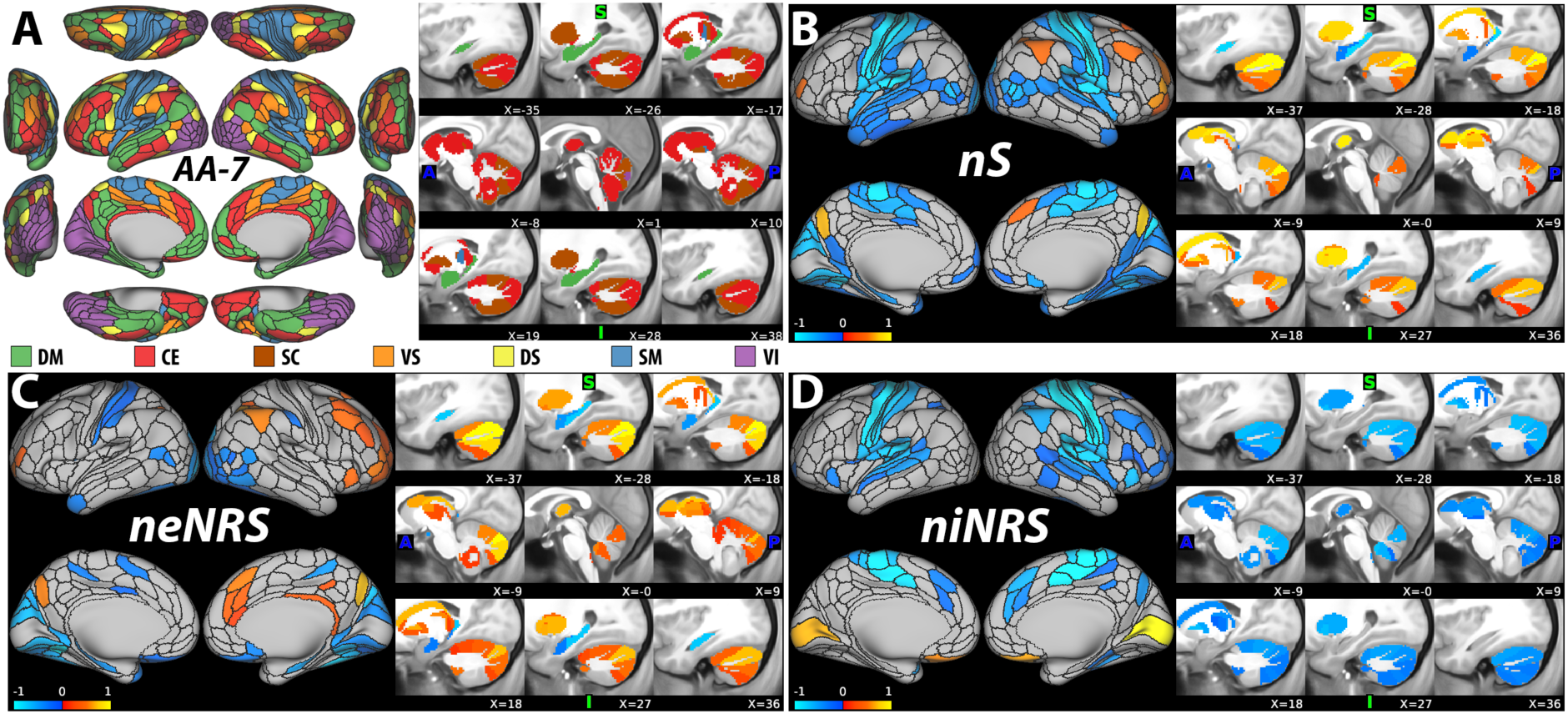
Ketamine Nodal Fingerprint (NFP). **A**. The nodal affiliation based on the Akiki-Abdallah (AA) hierarchical atlas at 7 canonical intrinsic connectivity networks (i.e., AA-7): default mode (DM), central executive (CE), subcortical (SC), ventral salience (VS), dorsal salience (DS), sensorimotor (SM) and visual (VI). The AA- 7 affiliation was used to compute nodal external network restricted strength (neNRS) and nodal internal NRS (niNRS). **B-D**. Nodal predictive results using nodal strength (nS; primary NFP; **B**), neNRS (**C**), or niNRS (**D**) as input features. Only nodes of significant predictive models following correction are shown (all *p* ≤ 0.001). The color bar unit is arbitrary, reflecting the sum of weighted nodes. All predictive models will be made publicly available at https://github.com/emergelab.

### Generalizability of the Ketamine Fingerprints

We next investigated whether the above fingerprints identified in Cohort A, hereafter termed Ketamine CFP/NFP, would generalize to new samples performing similar (Cohort B) or differing (Cohort C) tasks during infusion. Compared to normal saline, ketamine significantly increased the Ketamine CFP connectivity at AA-50, AA-24, and full connectome in both Cohort B and C (all *p* ≤ 0.001, *q* < 0.05; Fig. 3A). Ketamine also increased Ketamine NFP strength of niNRS and neNRS in both Cohort B and C (all *p* ≤ 0.01, *q* < 0.05; Fig. 3A). However, Ketamine NFP of nodal strength (nS) was significantly increased in Cohort C (*p* = 0.001, *q* < 0.05), but not Cohort B (*p* = 0.17; Fig. 3A).

**Figure 3.**
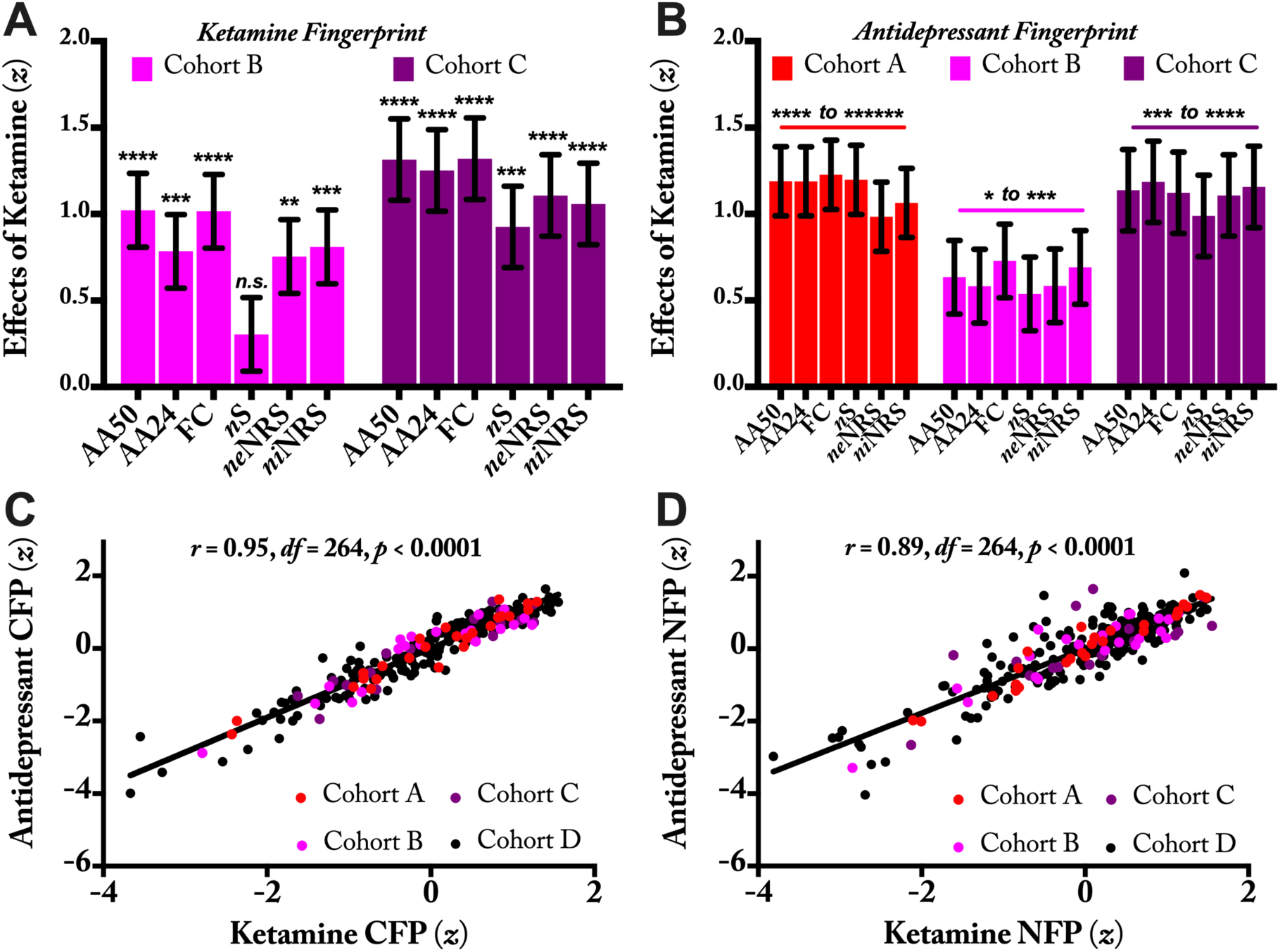
Reproducibility and Clinical Relevance of the Ketamine Connectome (CFPs) and Nodal Fingerprints (NFPs). **A**. The ketamine functional connectivity fingerprints identified in Cohort A were examined in Cohort B and C. Ketamine significantly increased all connectivity fingerprints during infusion, except the nodal strength (nS) NFP in Cohort B. **B**. In three cohorts of healthy participants, ketamine significantly increased connectivity of previously established antidepressant fingerprints. **C-D**. Across healthy and depressed subjects, there are high associations between the ketamine and antidepressant connectivity fingerprints. *Abbreviations* – AA: Akiki-Abdallah hierarchical connectivity atlas; AA50: CFP using AA at 50 modules; AA24: CFP using AA at 24 modules; FC: full connectome using 424 nodes (i.e., A424); neNRS: NFP using nodal external network restricted strength; niNRS: NFP using nodal internal NRS.

Considering the excellent generalizability of the Ketamine CFP, we further assessed the robustness and reproducibility of the NRS-PM in Cohort B and C. As shown in Fig. S3, NRS-PM analyses identified significant CFPs in both Cohort B and C, which appears to be qualitatively comparable to the Ketamine CFP established in Cohort A.

### Relevance to Antidepressants Treatment

To assess the relevance of the ketamine signatures to the effect of antidepressants, we first investigated the association between the connectivity of these fingerprints and the percent improvement in depression scores following treatment with sertraline, compared to placebo in Cohort D. We found a significant positive correlation between percent improvement in depression scores at Week 8 and increased connectivity at Week 1 in each of the Ketamine CFPs at AA-50 (*r* = 0.30, *df* = 199, *p* < 0.001), AA-24 (*r* = 0.27, *df* = 199, *p* < 0.001), and the full connectome (*r* = 0.29, *df* = 199, *p* < 0.001), and the Ketamine NFPs of nS (*r* = 0.21, *df* = 199, *p* = 0.003), niNRS (*r* = 0.21, *df* = 199, *p* = 0.003) and neNRS (*r* = 0.26, *df* = 199, *p* < 0.001).

Next, we investigated the effects of ketamine on the antidepressant fingerprints previously identified by Nemati, Akiki et al. [11]. As shown in Fig. 3B, we found that in each of Cohort A, B, and C, ketamine significantly increased connectivity in the Antidepressant CFPs and NFPs. Note that the data were not combined across cohorts in order to assess both the robustness (i.e., significance in small samples) and the reproducibility of the fingerprints across samples.

Finally, we investigated the association between the ketamine and antidepressant fingerprints. Across healthy and depressed subjects, we found positive correlations between Ketamine CFP and Antidepressant CFP (*r* = 0.95, *df* = 264, *p* < 0.001; Fig. 3C), as well as between Ketamine NFP and Antidepressant NFP (*r* = 0.89, *df* = 264, *p* < 0.001; Fig. 3D).

## DISCUSSION

The study successfully identified a robust and reproducible biomarker of ketamine’s effect on brain networks (i.e., Ketamine CFP). Secondary measures of CFPs were also significant. Both primary (AA-50) and secondary (AA-24 & full connectome) CFPs generalized to two separate samples (Cohort B & C). While the study identified a significant ketamine nodal fingerprint, this biosignature generalized to Cohort C, but not B, raising concerns about the reproducibility of the nodal strength (nS) biomarker. Nonetheless, it is important to note that as an average of all connections, nS is network agnostic and is not able to capture the dynamic shift within and between networks. As shown in Fig. 1, the Ketamine CFP is characterized by both reduction in internal and increase in external connectivity. For these network shifts, niNRS and neNRS are more suitable measures. As such, these secondary nodal fingerprints were highly robust and reproducible across all cohorts.

A major secondary finding is that the identified Ketamine CFP appears to be directly relevant to the treatment of depression, as evident by the positive correlation between the increase of Ketamine CFP connectivity and treatment response following antidepressant treatment compared to placebo in Cohort D. Moreover, ketamine was found to significantly increase Antidepressant CFP connectivity in each of Cohort A, B, and C. Furthermore, there is high association between the Ketamine and Antidepressant CFPs in both healthy and depressed populations.

Qualitatively, the Ketamine CFP may indicate a pattern of increased top-down control, showing a shift from internal connectivity within sensorimotor-visual and within central-executive-subcortical modules toward external connectivity between executive regions and primary cortices. In the default mode and ventral salience modules, there is also a shift from interference with primary cortices toward increased connection with the central-executive-subcortical modules (Fig. 1). This dynamic internal to external connectivity shift is also demonstrated quantitatively with the distribution of niNRS and neNRS nodal fingerprints. As evident in Fig. 2, nodes within the central executive network have shown both reduction in niNRS and increase in neNRS, which may have contributed to the dampened effects on nodal nS. The latter is a combination of internal and external connections (i.e., niNRS & neNRS).

The pattern of ketamine-induced enhanced executive connectivity is reminiscent of findings with the Antidepressant CFP [11]. It is also consistent with a model that associates depression with disconnection between the executive regions and the rest of the brain, combined with increased interference from the default mode and ventral salience modules, showing higher connections to primary cortices. In this model, ketamine and other antidepressants will exert their therapeutic effects by reversing this pathology. Of relevance, numerous MDD studies have shown reduced nS (also known as functional connectivity strength or global brain connectivity) within the prefrontal cortex (PFC) [13, 28-33], which might reflect the hypothesized executive disconnection in depression. Importantly, it was repeatedly shown that ketamine reverses this nS prefrontal dysconnectivity during infusion and at 24h post-treatment of depressed patients [13, 22, 28] (but not at 48h [30]). Other studies have also associated executive and default mode alterations with depression pathology and treatment [34-37].

The current results may relate to previous ketamine studies of functional connectivity [15, 38-42]. However, a limitation of the current method is that the modules are not directly comparable to previous seeds and networks. Furthermore, the Ketamine CFP was derived from task data collected during ketamine infusion and may differ from that observed under rest. Task-related connectivity typically reflects task cognitive operations and often differs from resting connectivity [43]. It has been shown that seed-based connectivity in the executive control network under ketamine differs between task and resting state [15]. Task data may have been especially useful, in the current investigation, because the oddball and the spatial working memory task increase demands upon the prefrontal cortex. Since depressed individuals have prominent prefrontal disruption [31], task data may highlight connectivity similarities between individuals on ketamine and depressed individuals.

Despite these limitations, the Ketamine CFP clearly shows that ketamine-induced connectivity changes combine increases and reductions in a network informed fashion. A leading model of the mechanisms of ketamine is that chronic stress pathology (CSP) and depression are associated with reduced synaptic connectivity (i.e., reduced synaptic density and strength) and that ketamine rapidly increases synaptic connectivity leading to robust and sustained antidepressant effects [44, 45]. In the PFC, and perhaps in the hippocampus, this CSP model has strong preclinical evidence and broad supportive human data [46]. However, what is often less highlighted in the literature is that CSP is also associated with increased synaptic connectivity in the nucleus accumbens and other brain regions [47, 48]. Conversely, ketamine effects are associated with reduced synaptic connectivity in the nucleus accumbens [47, 49]. Together, this data suggests that both increases and reductions in synaptic connectivity are required to successfully induce antidepressant effects. Considering that the ketamine-induced synaptic plasticity is activity-dependent [50], we hypothesize that during ketamine infusion, a glutamate surge [51] within the central executive synaptic connections gives rise to unique network shifts consistent with the functional connectivity changes captured by the Ketamine CFP. This transient pattern of altered synaptic neurotransmission is ultimately translated into changes of synaptic density and strength, which leads to rapid and sustained normalization of functional networks and to RAADs effects. This network-dependent model may provide partial explanation for the failure of other *N*-methyl-D-aspartate (NMDA) modulators [52]. For example, rapastinel has shown significant increase in PFC synaptic connectivity but failed in clinical trials [53]. Similarly, it may provide insight into why local injection of rapamycin into the PFC [54], but not its systemic administration [55, 56], block the ketamine RAAD effects. In this network-dependent model, it is hypothesized that interventions that affect synaptic connectivity regionally, without inducing the full spectrum of Ketamine CFP changes, are unlikely to induce robust antidepressant effects. Conversely, interventions that locally affect synaptic connectivity

## Data Availability

Data used in the preparation of this manuscript were obtained and analyzed from the controlled access datasets distributed from the NIMH-supported National Database for Clinical Trials (NDCT). NDCT is a collaborative informatics system created by the National Institute of Mental Health to provide a national resource to support and accelerate discovery related to clinical trial research in mental health. Dataset identifier(s): STU 092010-151; Establishing Moderators and Biosignatures of Antidepressant Response for Clinical Care (EMBARC).

## ACKNOWLEDGMENTS

The authors would like to thank the subjects who participated in these studies for their invaluable contribution. Data used in the preparation of this manuscript were obtained and analyzed from the controlled access datasets distributed from the NIMH-supported National Database for Clinical Trials (NDCT). NDCT is a collaborative informatics system created by the National Institute of Mental Health to provide a national resource to support and accelerate discovery related to clinical trial research in mental health. Dataset identifier(s): STU 092010-151; Establishing Moderators and Biosignatures of Antidepressant Response for Clinical Care (EMBARC). This manuscript reflects the views of the authors and may not reflect the opinions or views of the NIMH or of the individuals submitting original data to the NDCT.

This work was supported by National Center for Advancing Translational Science Grant No. 1UH2TR000960-01, National Institute on Alcohol Abuse and Alcoholism Grant No. P50AA12870, Yale Center for Clinical Investigation Grant No. UL1 RR024139, by NIMH (K23MH101498), GlaxoSmithKline, VA Career Development Award (IK2- CX0001873), American Foundation for Suicide Prevention (YIA-0- 004-16), and the Department of Veterans Affairs through its support for the Veterans Affairs National Center for Posttraumatic Stress Disorder. The content of this report is solely the responsibility of the authors and does not necessarily represent the official views of the sponsors, the Department of Veterans Affairs, NIH, or the U.S. Government.

(e.g., rapamycin in PFC), which subsequently disrupt the Ketamine CFP changes, may result in blockade of ketamine’s RAAD effects. However, it is critical to underscore the speculative nature of this network-dependent model and the need for confirmatory causal evidence in future studies.

## SUPPLEMENTAL INFORMATION

**Table S1.** Demographics and Clinical Characteristics of Cohort B.

|  | Sertraline (n = 97) | Placebo (n = 103) |
| --- | --- | --- |
| **Demographic** |  |  |
| Age, mean (SEM) | 38 (1.2) | 37 (12.2) |
| Women | 69% | 64% |
| Years of education, mean (SEM) | 15 (0.3) | 15 (0.2) |
| **Clinical features** |  |  |
| Age of onset, mean (SEM) | 14 (0.6) | 14 (0.6) |
| Baseline HAMD, mean (SD) | 19 (0.4) | 18 (0.4) |
| W1 HAMD, mean (SD) | 16 (0.6) | 16 (0.5) |
| W8 HAMD, mean (SD) | 10 (0.7) | 12 (0.7) |

\* There were no significant ( $p > 0.1$ ) differences between treatment groups. W1: week-1; W8: week-8; HAMD, Hamilton Rating Scale for Depression.

**Figure S1.**
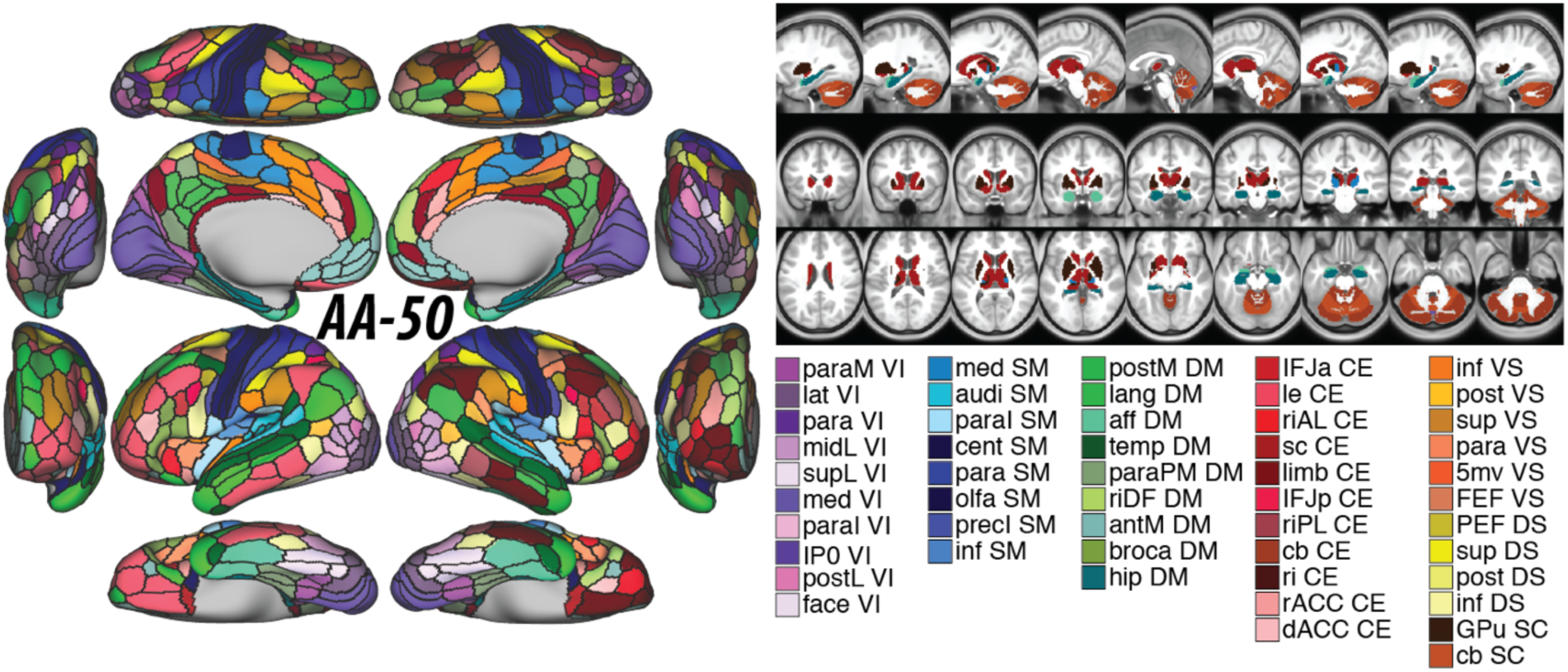
Whole-brain Akiki-Abdallah (AA) network affiliation at 50 modules architecture (i.e., AA-50). The module abbreviations of AA-50, along with further details about the affiliation of each node are publicly available at [1] and at https://github.com/emergelab/hierarchical-brain-networks/tree/master/brainmaps. The figure was adapted with permission from the Emerge Research Program (http://emerge.care).

**Figure S2.**
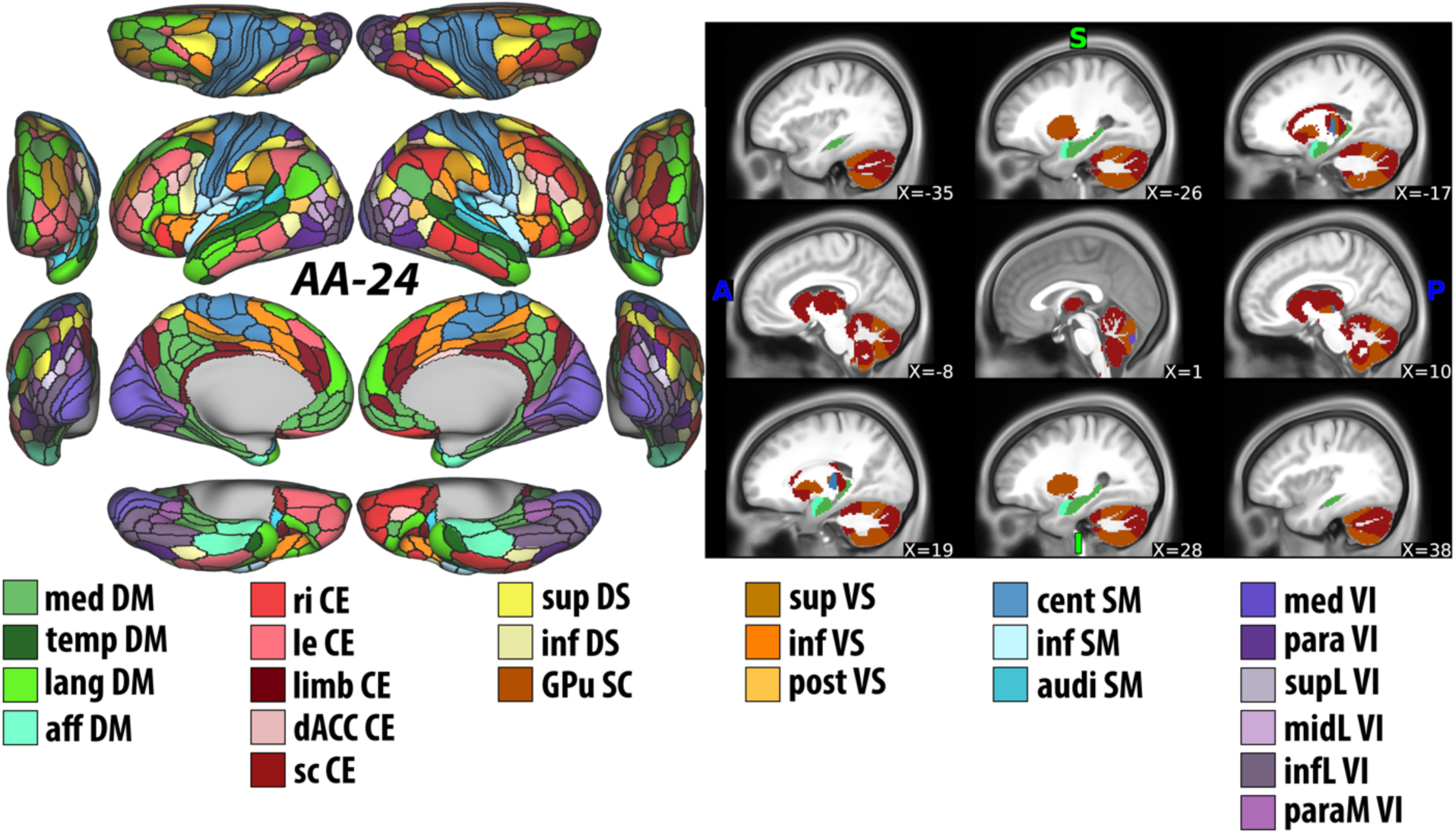
Whole-brain Akiki-Abdallah (AA) network affiliation at 24 modules architecture (i.e., AA-24). The module abbreviations of AA-24, along with further details about the affiliation of each node are publicly available at [1] and at https://github.com/emergelab/hierarchical-brain-networks/tree/master/brainmaps. The figure was adapted with permission from the Emerge Research Program (http://emerge.care).

**Figure S3.**
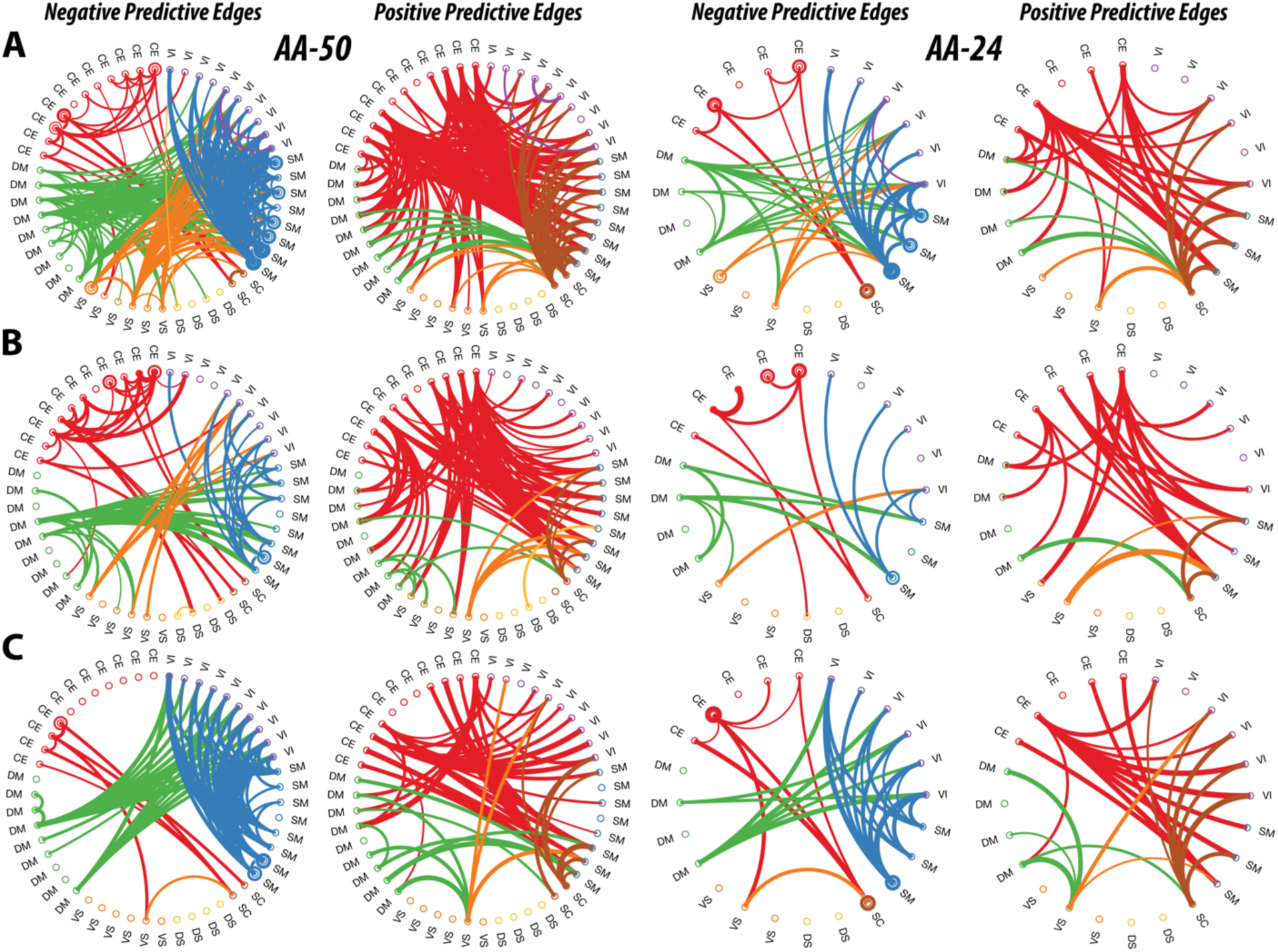
Ketamine Connectome Fingerprints in Cohort A, B, and C. The circular graphs are labeled based on the Akiki-Abdallah (AA) whole-brain architecture at 55 modules (AA-50; primary CFP), 24 modules (AA-24), and the full connectome with 424 nodes (A424). Modules and nodes are colored according to their affiliation to the 7 canonical connectivity networks: central executive (CE), default mode (DM), ventral salience (VS), dorsal salience (DS), subcortical (SC), sensorimotor (SM), and visual (VI). Edges are colored based on the initiating module using a counter-clockwise path starting at 12 o’clock. Internal edges (i.e., within module) are depicted as outer circles around the corresponding module. Thickness of edges reflect their corresponding weight in the predictive model. The module abbreviations of AA-24 and AA-50, along with further details about the affiliation of each node are available at https://github.com/emergelab. Only edges of significant predictive models following correction are shown (all *p* ≤ 0.002).

